# Can routine antenatal data be used to assess HIV antiretroviral therapy coverage among pregnant women? Evaluating the validity of different data sources in the Western Cape, South Africa

**DOI:** 10.1101/2023.09.08.23295270

**Authors:** Nisha Jacob, Brian Rice, Alexa Heekes, Leigh F. Johnson, Samantha Brinkmann, Tendesayi Kufa, Andrew Boulle

## Abstract

**Background:** Accurate measurement of antenatal antiretroviral treatment (ART) coverage in pregnancy is imperative in tracking progress towards elimination of vertical HIV transmission. In the Western Cape, South Africa, public-sector individual-level routine data are consolidated from multiple sources, enabling the description of temporal changes in population-wide antenatal antiretroviral coverage. We evaluated the validity of different methods for measuring ART coverage among pregnant women.

**Methods:** We compared self-reported ART data from a 2014 antenatal survey with laboratory assay data from a sub-sample within the survey population. Thereafter, we conducted a retrospective cohort analysis of all pregnancies consolidated in the Provincial Health Data Centre (PHDC) from January 2011 to December 2020. Evidence of antenatal and HIV care from electronic platforms were linked using a unique patient identifier. ART coverage estimates were triangulated with available antenatal survey estimates, aggregated programmatic data from registers recorded in the District Health Information System (DHIS) and Thembisa modelling estimates.

**Results:** Self-reported ART in the 2014 sentinel antenatal survey (n=1434) had high sensitivity (83.5%), specificity (94.5%) and agreement (k=0.8) with the gold standard of laboratory analysis of ART. Based on linked routine data, ART coverage by the time of delivery in mothers of live births increased from 67.4% in 2011 to 94.7% by 2019. This pattern of increasing antenatal ART coverage was also seen in the DHIS data, and estimated by the Thembisa model, but was less consistent in the antenatal survey data.

**Conclusion:** This study is the first in a high-burden HIV setting to compare sentinel ART surveillance data with consolidated individuated administrative data. Although self-report in survey conditions showed high validity, more recent data sources based on self-report and medical records may be uncertain with increasing ART coverage over time. Linked individuated data may offer a promising option for ART coverage estimation with greater granularity and efficiency.

## Introduction

Accurate measurement of antenatal antiretroviral treatment (ART) coverage in pregnancy is imperative in tracking progress towards the elimination of vertical transmission[1–3]. To monitor progress towards elimination of vertical transmission, the Global Plan for the elimination of mother-to-child transmission requires countries to report on three process measures with the targets that 95% of pregnant women attend antenatal care, 95% are tested for HIV and 95% of HIV positive pregnant women initiate ART prior to or during pregnancy[1].

Antenatal HIV sentinel surveillance is one approach to measuring these process indicators, and is a long-standing tool used in South Africa[4–7]. Since 1990, antenatal HIV sentinel surveys have taken place annually or biennially in all provinces of South Africa. Until 2015, the Western Cape province of South Africa extended the national survey to a larger proportionally weighted sample to generate sub-district level estimates. The World Health Organization (WHO) has recommended sentinel surveillance be replaced by routine programmatic data, a shift that is reliant on accurate and complete routine data[7,8]. While the focus of sentinel antenatal surveys is HIV seroprevalence, the widespread roll-out of ART necessitates the monitoring of ART coverage in conjunction with prevalence to accurately measure progress in managing HIV and preventing transmission[9].

In 2013, the Western Cape province of South Africa adopted a policy of lifelong ART for all pregnant women living with HIV (WLWH) (Option B+)[10,11]. Subsequently, the Western Cape Provincial Department of Health (WCDOH) added an additional question on ART use to the 2014 provincial antenatal survey, which was also incorporated in national surveys [12–14]. This question measured current ART use in the 3 days prior to survey participation[12]. The validity of self-reported ART use as a method to evaluate ART coverage using existing sentinel surveillance methods requires formal evaluation. An alternative method for measuring ART use is using routine data. The WCDOH makes use of two routine health information platforms. Firstly, aggregated data from facility-based registers are captured in the District Health Information System (DHIS). Secondly, individuated electronic patient data are consolidated by the Provincial Health Data Centre (PHDC). These include administrative, pharmacy and laboratory data linked via a unique patient folder number, in the absence of electronic medical records. ART coverage can therefore be estimated using pharmacy dispensing data of triple-regimen ART or entries in HIV treatment registers.

We evaluated the validity of different methods for measurement of ART coverage by assessing self-reported ART use in sentinel antenatal surveys and comparing sentinel survey estimates with routine programmatic data estimates over a 10-year period.

## Methods

This study was conducted in the Western Cape province of South Africa. The province is comprised of six districts: Cape Metro, Overberg, Garden Route, Central Karoo, West Coast and Cape Winelands. The study was comprised of two parts: validation of 2014 self-reported ART data with laboratory data, and the comparison of cross-sectional sentinel survey ART coverage estimates as well as modelling estimates with routine data from 2011 to 2020. The datasets used are described below.

### Antenatal survey data

Antenatal ART coverage estimates for the Western Cape Province for 2014 and 2015 were obtained from the Western Cape provincial sentinel antenatal survey. In 2016, the provincial survey was discontinued and annual national surveys were replaced by biennial surveys. Antenatal ART coverage estimates for the Western Cape province for 2017 and 2019 were obtained from the South African National Department of Health as reported in the national sentinel antenatal survey reports.

Estimates were derived from data that included all pregnant women attending their first antenatal visit in a public health facility during a 6-week period of a survey in years 2014 and 2015. In years 2017 and 2019, the national survey data included pregnant women attending first antenatal visits or follow-up antenatal visits in a public health facility during a 6-week period (in this current analysis, only data collected at first antenatal visit are included). The survey question for current ART use in 2014, 2015 and 2017 surveys was: “Have you taken ARVs in the last 3 days?”. This question measured ART use at enrolment as a proxy for ART use prior to the pregnancy or by the time of conception, excluding any prior treatment interruption. This question was discontinued in 2019. In 2017 and 2019, participants were asked if they had ever taken ARVs and if so when, therefore measuring any ART use (prior or current). From 2019, the health care worker asking the question was further expected to verify the response with information in the participant’s medical records if available.

### Laboratory validation sub-sample

As part of a WCDOH analysis, a third of the HIV positive specimens from the 2014 provincial antenatal survey were sampled, stratified by district, obtaining a sub-sample of 450 specimens. These were tested by the University of Cape Town (UCT) Department of Pharmacology for lopinavir, efavirenz and nevirapine, using a cut-off of 50ng/ml. At least one of these drugs were included in all ART regimens in use in South Africa in 2014. These data were used to validate the additional survey question on current ART use introduced in the 2014 provincial antenatal survey.

### Provincial Health Data Centre (Routine individual-level HIV program data)

A retrospective cohort of pregnancies was enumerated from the PHDC on 26 May 2022, which included anonymised linked data of all pregnant women attending public health facilities in the Western Cape between 2011 to 2020. Pregnancies were allocated a quantitative confidence score to measure the degree of confidence in the recorded information/data points, indicating a true pregnancy. Pregnancies scoring 0.7 or higher out of a maximum of 1 were included as they have at least 1 high confidence data point such as a rhesus antibody test (conducted routinely at first antenatal visit), pregnancy test, International Classification of Diseases (ICD) Tenth Revision code indicating pregnancy, antenatal visit or maternal discharge summary. The geographic location of the facility of first pregnancy-related attendance was used to determine district of pregnancy. Pregnancy outcome date and available information on gestational age were used to estimate pregnancy period, with the date of the first evidence of pregnancy used to stratify pregnancies by year. Within this cohort, administrative, laboratory and pharmacy evidence of HIV diagnosis before or during the estimated pregnancy period was used to determine antenatal HIV status. Antenatal ART coverage was based on the proportion of women known to be living with HIV during the antenatal period with evidence of ART initiation before or during pregnancy. Women who interrupted treatment would be included as having started ART by enrolment.

### District Health Information System (Routine aggregated HIV program data)

Aggregated program data of women recorded as attending public antenatal facilities in the Western Cape for the first time in 2011 to 2020 were included in the DHIS dataset. For this analysis, the element “Known HIV positive client on ART at first visit” was used to determine ART coverage at first antenatal visit among all those known with HIV before pregnancy and diagnosed with HIV at first antenatal visit. This data element is intended to measure ART use at conception and excludes women who had interrupted treatment at the time of the first antenatal visit.

### Thembisa Model

Thembisa is an integrated demographic and HIV model, developed for South Africa. The model is applied at both national and provincial levels; the estimates used in this study are from version 4.6 of the Western Cape model[15]. The provincial model is calibrated to a number of HIV data sources: age-specific HIV prevalence data from antenatal clinic surveys and household surveys, recorded numbers of deaths by age and sex, total numbers of ART patients and age/sex distributions of ART patients, and antiretroviral metabolite data. Assumptions about fertility rates in HIV-positive women and the effect of ART on fertility are informed by Western Cape PHDC data[16]. The model makes assumptions about rates of ART interruption and re-initiation, which make it possible to estimate both current ART coverage and proportions of WLWH ever initiated on ART at the time of conception.

### Analysis

Data were analysed using Stata 17 (Stata Corporation, College Station, Texas, USA). Descriptive characteristics of the laboratory validation sub-sample were tabulated. Given the higher validity of laboratory-based ART testing[17], this dataset functioned as the gold standard for comparison with the newly introduced survey question in 2014[9]. Discordance between self-report and laboratory ART was calculated. Demographic risk factors for discordance were analysed using logistic regression. Sensitivity, specificity, positive predictive value and negative predictive value were calculated to validate the survey question on ART use. Agreement between self-report and lab ART detection was quantified using the kappa statistic.

Given the 10-year duration of the PHDC cohort (2011 – 2020) and the notable changes in ART guidelines and data availability over this time period, descriptive characteristics of the cohort were tabulated across all years and for the 2019 cohort specifically, as the most recent year pre-COVID19, to reflect the most recent characteristics.

ART-specific survey data and time trends from 2014 to 2019 were compared with PHDC, DHIS and Thembisa 4.6 estimates. The 2014 and 2015 provincial antenatal survey measured current ART use (excluding treatment interruption). The 2017 and 2019 national antenatal surveys measured any ART use before pregnancy (which may include treatment interruption). ART coverage estimates from the surveys were compared with PHDC estimates for women initiating ART before pregnancy among those with prenatal or antenatal evidence of HIV diagnosis (includes treatment interruption). The survey data were similarly compared with DHIS data which were limited to first visit ART coverage (excluding treatment interruption). The survey estimates, PHDC ART coverage estimates before pregnancy and DHIS estimates were further compared with ART at conception estimates from the Thembisa mathematical model estimates for the Western Cape, with and without treatment interruption [15].

PHDC estimates for women initiating ART before or during pregnancy among those with prenatal or antenatal evidence of HIV diagnosis (both with live births or any pregnancy outcome) were further compared with Thembisa estimates of ART at delivery among those with live births. A sensitivity analysis including moderate-low confidence pregnancies was conducted to determine the contribution of these pregnancies to ART coverage. The PHDC data were analysed further for estimates of ART coverage by district and age-group. These analyses were limited to a 5-year period (2015 – 2020) when the test and treat ART policy was fully implemented and individuated electronic data systems were better established.

### Ethical considerations

The study was approved by the University of Cape Town Human Research Ethics Committee (HREC 083/2021) and the Western Cape Provincial Health Research Committee. All antenatal HIV survey data, DHIS and Thembisa data were provided in aggregated form. Data from the PHDC were de-identified before study release according to the Western Cape Department of Health Data Access Policy Guidelines.

## Results

From 1 January 2011 to 31 December 2020, 977 800 and 989 568 pregnancies were recorded by the PHDC and DHIS, respectively, with the PHDC enumerating more pregnancies than DHIS from 2015 onwards. Sample sizes for the provincial surveys in 2014 and 2015 were 7470 and 7517, respectively. Following the discontinuation of provincial surveys, the Western Cape sample size of the national surveys in 2017 and 2019 were 3571 and 3943, respectively.

The descriptive characteristics of the laboratory sub-sample were comparable to the 2014 HIV positive survey population (Table 1). Within the sub-sample, 13 participants (2.9%) did not disclose ART status.

**Table 1:**
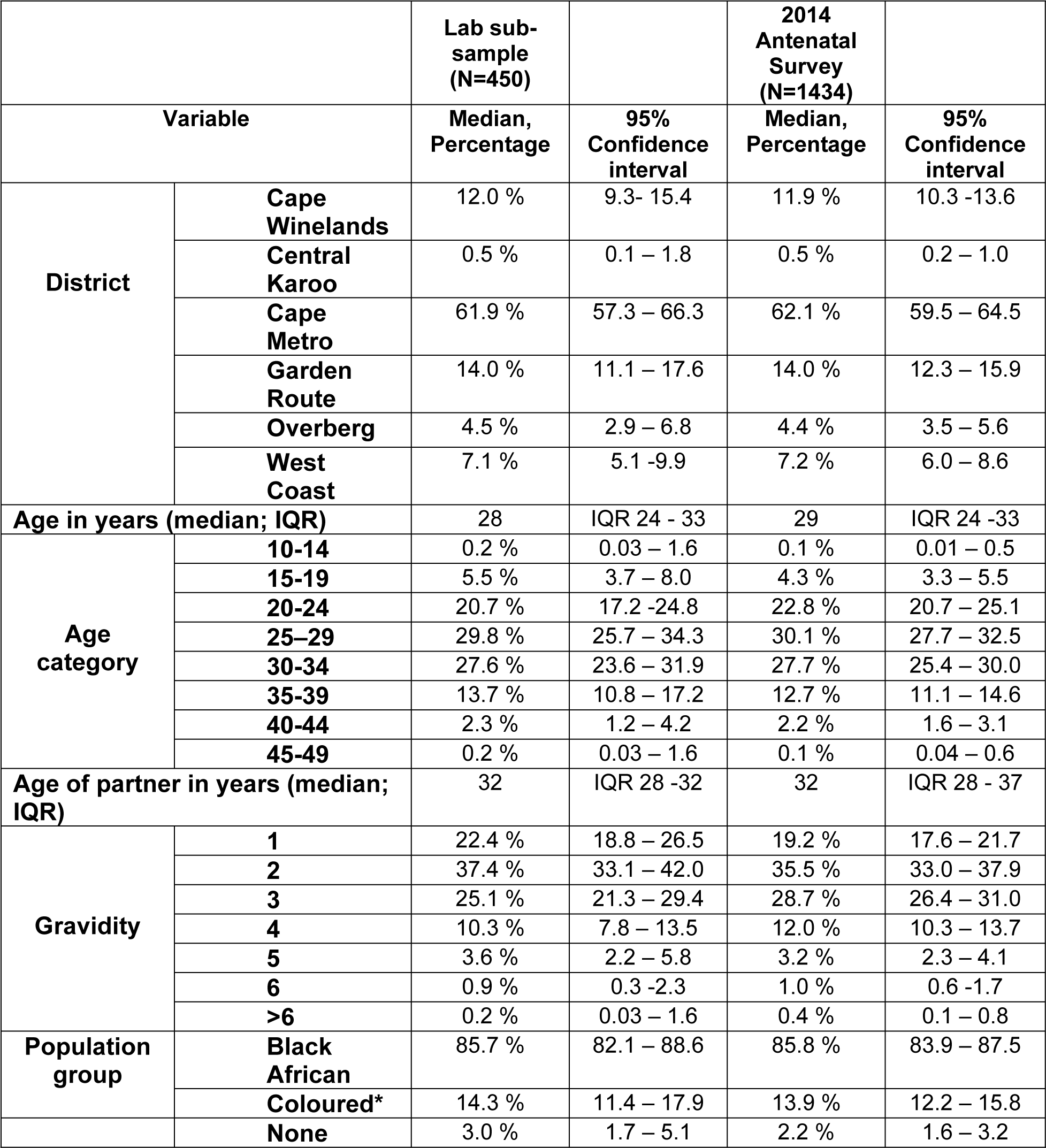
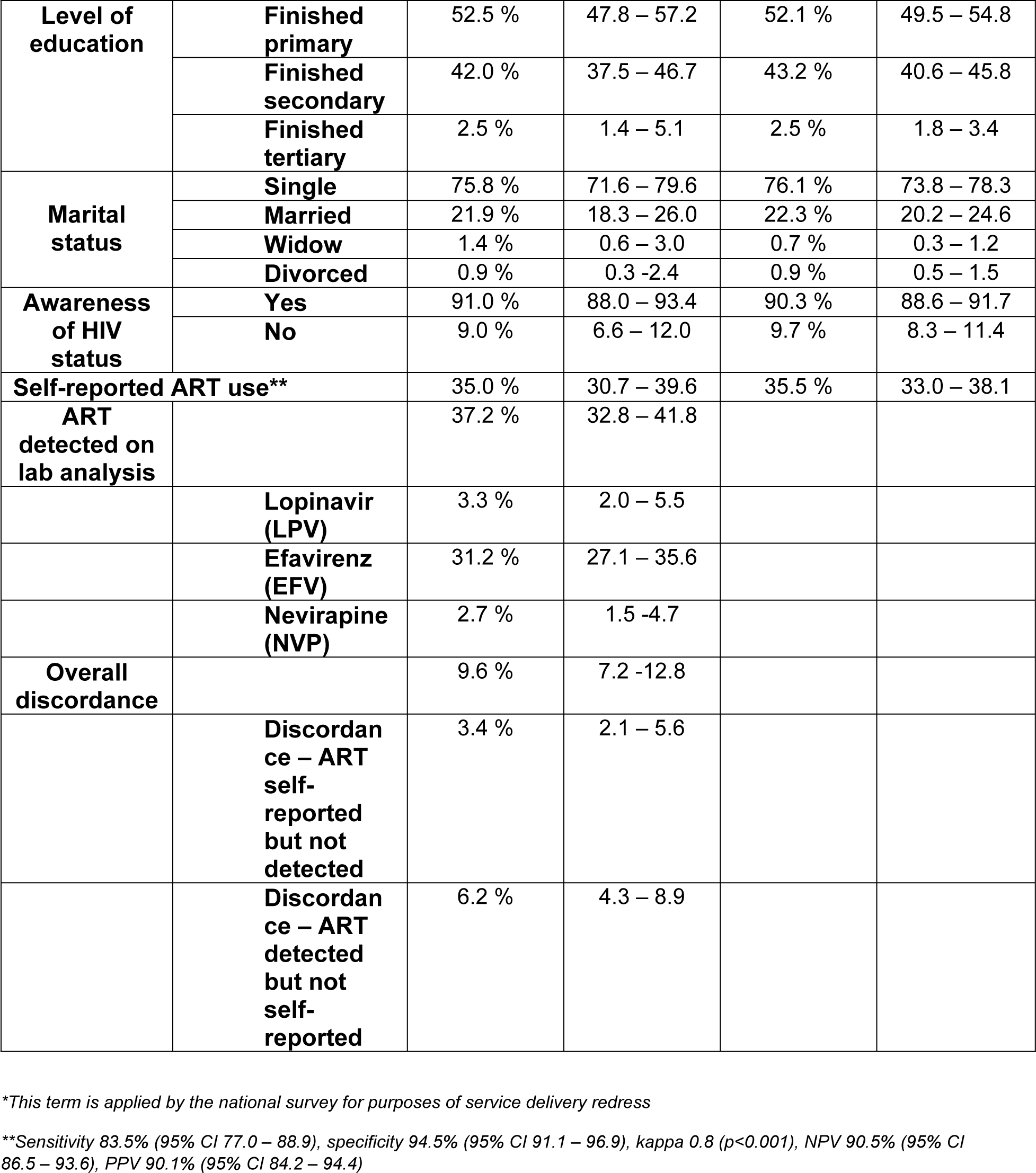
Characteristics of HIV-positive women in the laboratory validation sub-sample compared to antenatal survey, 2014.

No risk factors for discordance between the laboratory sub-sample and 2014 survey population were noted on bivariable and multivariable analyses. Self-reported ART use (35%) was similar to ART detected on lab analysis (37.2%), with 2.2% absolute difference. Self-reported ART was further compared to laboratory markers showing a sensitivity of 83.5% (95% CI 77.0 – 88.9) and a specificity of 94.5% (95% CI 91.1 – 96.9), positive predictive value of 90.1% (95% CI 84.2 – 94.4) and negative predictive value of 90.5% (95% CI 86.5 – 93.6). Agreement between self-report and laboratory markers was also high at 90.4% (kappa statistic 0.8, p<0.001).

Descriptive characteristics of the PHDC cohort are summarised in Table 2. The majority of women (69.8%) having no electronic record of HIV status, suggests either HIV negative (most commonly given high antenatal HIV testing coverage) or unknown status. Of those with electronic records of positive HIV status, 7.7% had no record of ART (Table 2). Of all pregnant women with HIV in the 10-year cohort, 72.6% (95% CI 72.4 – 72.8) had records of ART initiation before or during pregnancy and 87.2% (95% CI 86.8 – 87.6) in 2019.

**Table 2:** Descriptive and HIV-specific characteristics of PHDC Cohort (2011 – 2020) compared to PHDC Cohort 2019.

|  | PHDC Cohort 2011 – 2020 (n=977800) Percentage | 95% CI | PHDC Cohort 2019<sup>s</sup> (n=123907) Percentage | 95% CI |
| --- | --- | --- | --- | --- |
| **Prior electronic evidence of pregnancy\*** |  |  |  |  |
| 1 | 60.3% | 60.2 – 60.4 | 52.4% | 52.1 – 52.6 |
| 2 | 27.2% | 27.1 – 27.3 | 29.7% | 29.5 – 30.0 |
| 3 | 9.2% | 9.2 – 9.3 | 12.4% | 12.2 – 12.6 |
| 4 | 2.5% | 2.48 – 2.54 | 4.0% | 3.9 – 4.1 |
| 5 or more | 0.8% | 0.8 – 0.9 | 1.5% | 1.4 – 1.6 |
| **Age (median; IQR)** | 27.4 | 22.5 – 31.8 | 27.2 | 22.8 – 32.3 |
| **Age Category** |  |  |  |  |
| <15 | 0.5% | 0.4 – 0.5 | 0.5% | 0.4 – 0.5 |
| 15 -19 | 12.0% | 12.0 – 12.1 | 11.1% | 10.9 – 11.3 |
| 20 - 24 | 26.6% | 26.5 – 26.7 | 25.7% | 25.5 – 26.0 |
| 25 - 29 | 27.4% | 27.3 – 27.5 | 27.3% | 27.1 – 27.6 |
| 30 - 34 | 20.1% | 20.1 – 20.2 | 20.8% | 20.6 – 21.0 |
| 35 - 39 | 10.4% | 10.3 – 10.4 | 11.5% | 11.3 – 11.7 |
| >39 | 3.0% | 2.97 – 3.03 | 3.1% | 3.0 – 3.2 |
| **District\*\*** |  |  |  |  |
| Cape Winelands | 13.4% | 13.3 – 13.5 | 13.1% | 12.9 – 13.2 |
| Central Karoo | 0.8% | 0.7 – 0.8 | 1.2% | 1.1 – 1.3 |
| Cape Metro | 70.2% | 70.1 – 70.3 | 65.7% | 65.4 – 65.9 |
| Garden Route | 8.3% | 8.3 – 8.4 | 9.2% | 9.0 – 9.4 |
| Overberg | 3.0% | 2.9 – 3.0 | 4.3% | 4.2 – 4.4 |
| West Coast | 3.7% | 3.7 – 3.8 | 5.5% | 5.4 – 5.6 |
| No district recorded | 0.6% | 0.61 – 0.64 | 1.1% | 1.0 – 1.1 |
| **Pregnancy Outcomes** |  |  |  |  |
| Delivered | 83.4% | 83.3 – 83.5 | 81.1% | 80.1 – 81.3 |
| Ectopic pregnancy | 1.4% | 1.4 – 1.5 | 1.7% | 1.7 – 1.8 |
| Termination | 5.4% | 5.3 – 5.4 | 7.2% | 7.1 – 7.4 |
| Stillbirth | 1.5% | 1.4 – 1.5 | 1.3% | 1.2 – 1.4 |
| Miscarriage | 3.4% | 3.4 – 3.5 | 4.9% | 4.8 – 5.0 |
| Mother died prior to delivery | 0.02% | 0.01 – 0.02 | 0.02% | 0.01 – 0.03 |
| Outcome unknown | 4.9% | 4.86 – 4.94 | 3.8% | 3.7 – 3.9 |
| **Evidence of HIV status** |  |  |  |  |
| HIV status unknown† | 69.8% | 69.8 – 69.9 | 71.6% | 71.6 – 72.1 |
| HIV status known | 30.2% | 30.1 – 30.3 | 27.9% | 27.9 – 28.4 |
| **HIV evidence timing** |  |  |  |  |
| HIV positive before or during pregnancy | 16.7% | 16.7 – 16.8 | 18.5% | 18.3 – 18.7 |
| HIV positive postnatally | 4.0% | 4.00 – 4.03 | 1.4% | 1.3 – 1.5 |
| HIV negative | 9.5% | 9.4 – 9.5 | 8.3% | 8.1 – 8.4 |
| HIV unknown | 69.8% | 69.7 – 69.9 | 71.8% | 71.6 – 72.1 |
| **ART (n=202575)** |  |  | **(n=24644)** |  |
| HIV Positive but no evidence of ART | 7.7% | 7.6 – 7.8 | 5.9% | 5.6 – 6.2 |
| Antenatal ART (before or during pregnancy) | 72.6% | 72.4 – 72.8 | 87.2% | 86.8 – 87.6 |
| Postnatal ART | 19.7% | 19.6 – 19.9 | 6.9% | 6.6 – 7.2 |
| **Antenatal ART (n=163649)‡** |  |  | **(n=22910)** |  |
| ART started before pregnancy | 54.7% | 54.4 – 54.9 | 69.5% | 68.9 – 70.1 |
| ART started during pregnancy | 35.2% | 35.0 – 35.4 | 24.3% | 23.7 – 24.9 |
| No antenatal ART | 10.1% | 10.0 – 10.3 | 6.2% | 5.9 – 6.5 |

§2019 reflects the most recent characteristics pre COVID-19
\*Gravidity estimates in the PHDC are not reliable since historic data are incomplete. Prior electronic evidence is used as a proxy to provide a full description of the cohort.
\*\*Unweighted
†HIV diagnosis is based on point-of-care tests that are not recorded electronically. Unknown HIV status therefore includes those who tested negative on point-of-care tests and therefore have no subsequent electronic evidence of HIV
‡Excludes women who only tested HIV positive postnatally

Among those living with HIV, survey, DHIS, PHDC and Thembisa estimates show that ART coverage before pregnancy increased over time, however a slight decrease is noted in the survey estimate in 2019 (Table 3 and Figure 1). The 2014 laboratory and PHDC estimates of ART coverage were almost the same (37.2% v 37.4%, respectively). The 2015 survey estimate of 44.2% was similar to the PHDC estimate of 42.8%. The 2017 survey estimate of 62.3% was higher than PHDC data (58.4%), but lower in 2019 (61.2%) compared to PHDC (69.5%), when comparing ART coverage before pregnancy. DHIS estimates were notably higher than Thembisa estimates, excluding treatment interruption, and more aligned to PHDC estimates which included treatment interruption. Thembisa estimates of those ever on ART prior to conception (including treatment interruption), were higher than PHDC until 2016 and thereafter reasonably close to PHDC estimates. From 2015 to 2020, Thembisa estimates of ART at delivery were higher than PHDC estimates of ART at delivery, however the difference between estimates is lower when restricting PHDC estimates to live births only. A sensitivity analysis to assess the contribution of moderate-low confidence pregnancies to ART coverage did not materially impact the PHDC estimates and associated comparisons.

**Figure 1:**
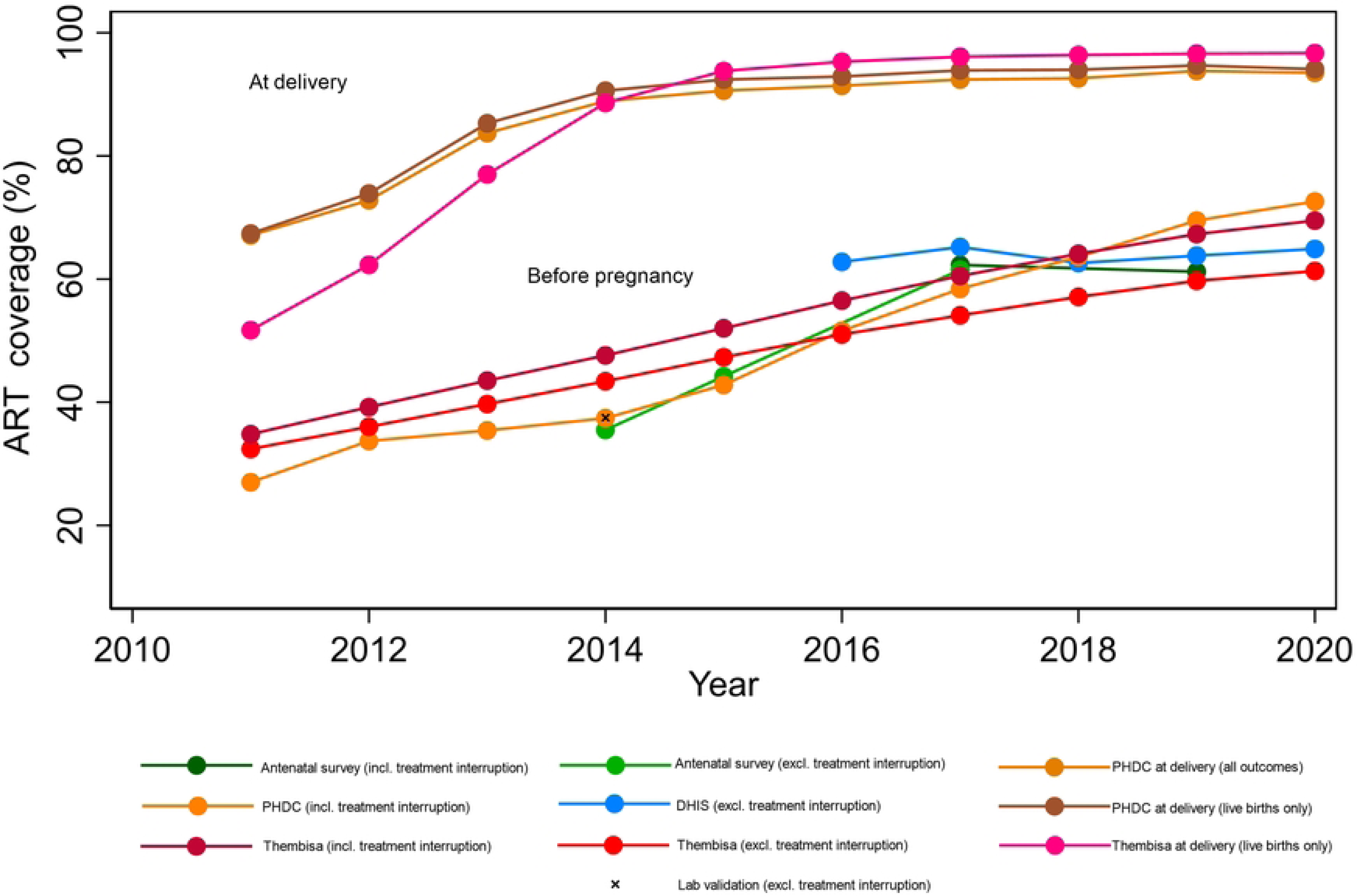
ART coverage before and during pregnancy by dataset (2011 – 2020)

**Table 3:** Western Cape Antenatal ART Coverage (%) amongst women living with HIV by dataset (2011 – 2020)

|  | Before pregnancy |  |  |  |  |  | At delivery |  |  |
| --- | --- | --- | --- | --- | --- | --- | --- | --- | --- |
|  | Antenatal Survey |  | DHIS | Thembisa 4.6 | PHDC | Thembisa 4.6 | PHDC | PHDC | Thembisa 4.6 |
| Year | Antenatal Survey (on ART among WLWH) | Lab validation | Proportion on ART among those with HIV at antenatal first visit (excluding treatment interruption) | Antenatal ART coverage prior to conception (excluding treatment interruption) | Proportion on ART before pregnancy among those known with HIV (including treatment interruption) | Antenatal ART coverage prior to conception (including treatment interruption) | Antenatal ART coverage at delivery (all pregnancy outcomes) | Antenatal ART coverage at delivery (live births only) | Antenatal ART coverage at delivery (live births only) |
| 2011 |  |  |  | 32.4 | 27.0 | 34.8 | 67.1 | 67.4 | 51.7 |
| 2012 |  |  |  | 36.0 | 33.7 | 39.2 | 72.8 | 73.9 | 62.3 |
| 2013 |  |  |  | 39.7 | 35.4 | 43.5 | 83.7 | 85.3 | 77.0 |
| 2014 | 35.5 (33.0 - 38.0)* | 37.2 (32.7 - 41.7)* |  | 43.4 | 37.4 | 47.6 | 88.9 | 90.6 | 88.6 |
| 2015 | 44.2 (41.6 - 46.8)* |  | 43.9 | 47.3 | 42.8 | 52.0 | 90.6 | 92.4 | 93.8 |
| 2016 |  |  | 51.4 | 51.0 | 51.6 | 56.5 | 91.4 | 92.9 | 95.3 |
| 2017 | 61.5 (57.7 - 65.1)*<br>62.3 (58.5 - 66.0)** |  | 62.0 | 54.1 | 58.4 | 60.5 | 92.4 | 93.9 | 96.1 |
| 2018 |  |  | 62.9 | 57.1 | 63.6 | 64.1 | 92.6 | 94.0 | 96.4 |
| 2019 | 61.2 (57.2 - 65.1)** |  | 68.3 | 59.7 | 69.5 | 67.3 | 93.8 | 94.7 | 96.6 |
| 2020 |  |  | 70.1 | 61.3 | 72.6 | 69.5 | 93.5 | 94.1 | 96.7 |

\*Current ART use (excludes treatment interruption)
\*\* Includes treatment interruption. Stratified to first visit only. ART before pregnancy amongst first and follow-up attendees is 58.4 for 2017 and 58.2 for 2019.

Figure 2 presents Western Cape antenatal ART coverage among those with HIV over a 10-year period according to PHDC data. ART coverage prior to pregnancy amongst women living with HIV increased over time from 42.8% (95% CI 42.0 - 43.5) in 2015 to 72.6% (95% CI 72.0 - 73.1) in 2020. Higher coverage was observed in older age groups, with coverage as high as 81.1% in the 35-39 age group in 2020 (Figure 1). An increasing proportion of women are shown to be on ART before pregnancy and from 2015 onwards ART coverage exceeds 90% before or during pregnancy. ART coverage during the antenatal period (ART commenced before or during pregnancy) was high amongst pregnant WLWH in all districts, except Central Karoo, and across all years. Over time, there was variability of coverage estimates in Central Karoo, Overberg and West Coast, with Cape Metro having the most consistent increase in coverage.

**Figure 2:**
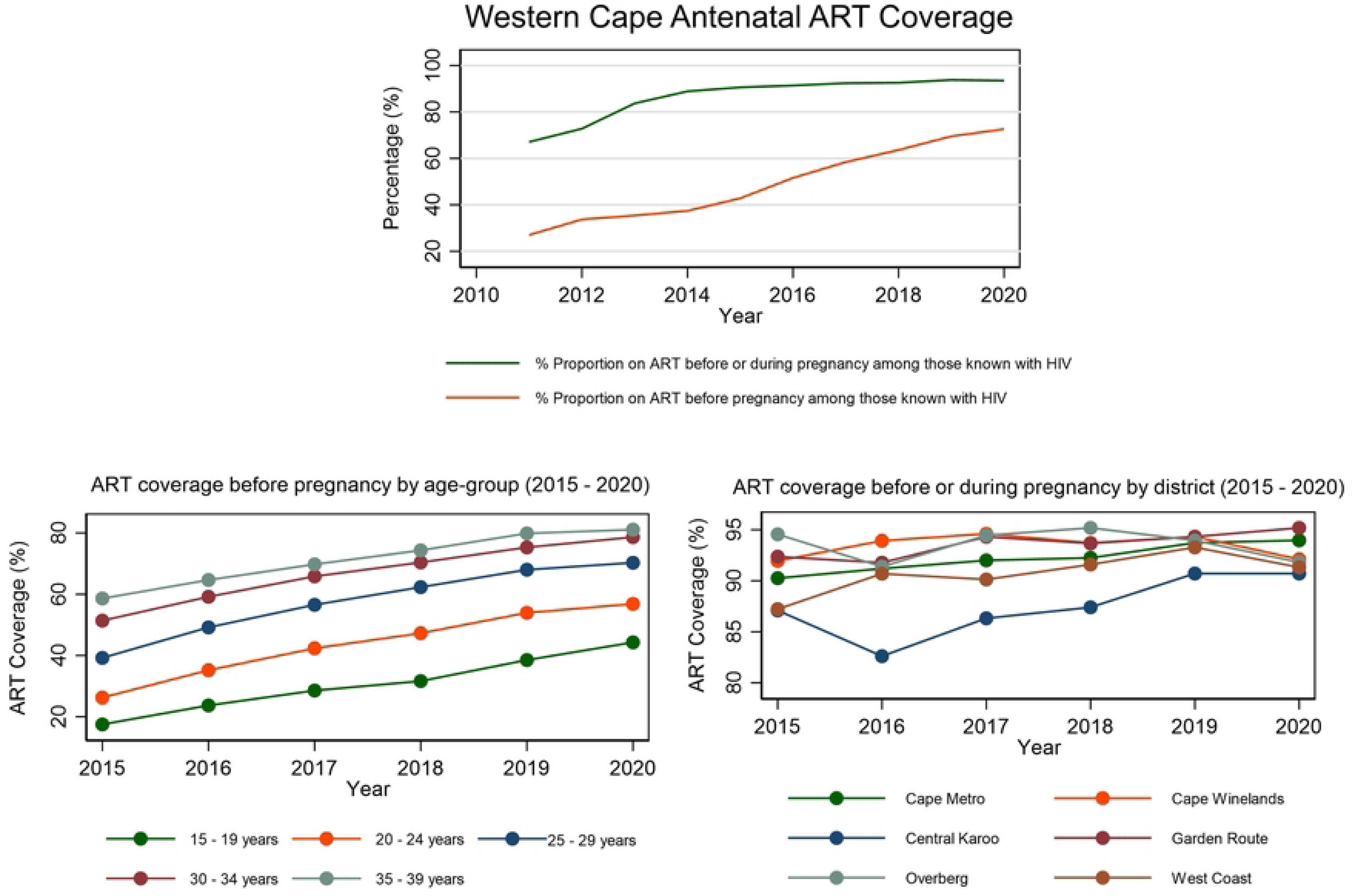
Temporal trends in antenatal ART coverage in the Western Cape by timing, district and age (PHDC)

## Discussion

Our study is the first to assess and compare different methods of antenatal ART coverage ascertainment in the Western Cape province. All datasets documented the increasing ART coverage in pregnant women over time. The challenges in dataset comparisons where numerators and denominators are disparate were highlighted.

The self-reported survey-based antenatal ART question was confirmed as a valid measure of ART use by comparison to the gold standard of laboratory detection, with good overall concordance. Discordance was higher amongst those self-reporting no ART use but having positive laboratory detection of ART than vice versa. This may represent misunderstanding of the question, false positive laboratory results or reluctance in disclosing ART use due to stigma or other reasons. Interestingly, 15 participants who self-reported ART use had no laboratory evidence of ART. In addition to question misinterpretation or false negative laboratory results, this may also suggest poor recent medication adherence. Discordance proportions and agreement between self-report and laboratory detection were in keeping with a similar study conducted in the Kwazulu-Natal province, however non-disclosure of ART status (2.9%) was notably lower in our cohort compared to this study[9]. Several other studies in sub-Saharan Africa comparing self-report and biomarkers of current or previous ART use showed higher proportions of ART non-disclosure, however these studies had smaller sample sizes and the study population was not restricted to pregnant women presenting for antenatal care[18–21]. While the study by Huerga et. al. in Kwazulu Natal found younger individuals were at higher risk of discordance, our study found no demographic risk factors for discordance[9]. The ART question had high agreement beyond chance, and high specificity, sensitivity and positive and negative predictive values, suggesting good performance of this question as a measure of ART use. Other studies have shown a similarly high specificity and positive predictive value, however sensitivity and negative predictive value are notably lower[22–24]. Reasons for better performance of self-reported ART in our study may include lower social desirability bias due to lower perceived stigma of ART use amongst those actively seeking antenatal care in a clinical setting. Furthermore, since the ART question was asked by clinicians during routine clinical care, more explanation of the question may have been given compared to other study settings. Our findings arguably support the continued use of the question in survey settings and the utility of survey ART estimates for further validation with routine data. However, we note that the question on current ART use was discontinued after 2017 and replaced with a question “has the participant ever taken ARV’s,” thereby including those not on ART at the time due to prior treatment interruption. Additionally, from 2019, survey questions were complemented with information from medical records if available. Further validation of survey data is therefore recommended.

From 2015 onwards, the PHDC enumerated more pregnancies than DHIS, suggesting good completeness of these data. Higher numbers in the PHDC may be due to inclusion of non-viable pregnancies and terminations of pregnancy, which may not be captured in DHIS amongst antenatal service attendees. Lower enumeration prior to 2015 is likely due to less well-established electronic systems in earlier years. Within the PHDC cohort, 69.8% had no information of HIV status. This is likely representative of the HIV negative antenatal population whose HIV status is determined solely by point-of-care rapid HIV testing, the results of which are not captured electronically on routine information systems [25]. Those with evidence of HIV negative status had laboratory confirmation via HIV ELISA test results which are captured electronically on routine systems. Those testing positive for HIV on point-of-care testing may have further validation of test results by laboratory HIV ELISA testing as well as other laboratory, pharmacy and administrative evidences of HIV, thus providing a complete dataset of HIV positive antenatal patients in the public sector of the Western Cape province [25].

Estimates of ART coverage in the HIV positive antenatal population were similar across the survey and PHDC datasets for 2014 and 2015, although higher estimates were expected in 2015 due to inclusion of treatment interruption in the PHDC dataset. The 2017 and 2019 antenatal surveys report ART use prior to pregnancy among women presenting for their first antenatal visit, and therefore may include those who interrupted treatment [13,14]. When compared to PHDC estimates of ART before pregnancy, PHDC estimates were lower than survey estimates in 2017, but notably higher in 2019. The differences between survey estimates and PHDC may reflect sampling bias in the survey population as well as interviewer-related differences in asking the survey question on ART use prior to pregnancy, participant-related interpretation of the question as well as availability of relevant medical records in 2019 survey participants. Whether the estimates include or exclude treatment interruption may be dependent on how the survey question was interpreted by both interviewer and participant.

Thembisa estimates of ART at delivery were higher than PHDC estimates, however the difference was slightly less when restricting PHDC estimates to live births only. This may indicate that ART initiation is prioritised where transmission to an infant is possible, and less urgent in non-viable pregnancies such as ectopic pregnancies. Additionally, pregnancies where the outcome is unknown may represent those who are lost to the health care system for reasons such as migration to a different province and therefore not recorded on ART within the PHDC. The high ART coverage estimates reflected in the PHDC dataset is reassuring and in keeping with recent literature[26]. These findings support the importance of antenatal first visits for early engagement in HIV care and effective prevention of vertical transmission of HIV. Any antenatal engagement with health services for WLWH, regardless of pregnancy intention or outcome, should be considered an opportunity for ART initiation in keeping with test and treat guidelines. Systemic factors associated with delayed ART initiation requires urgent research in order to optimise prevention of vertical transmission of HIV.

DHIS estimates of ART coverage among WLWH at first antenatal visit are notably higher than survey estimates and Thembisa estimates and more aligned to PHDC estimates, despite differences in treatment interruption ascertainment. The reasons for these disparities may be multifactorial and difficult to identify. It should be noted that Thembisa estimates include both the private and public sector. When including treatment interruption, Thembisa estimates are notably higher than PHDC estimates until 2016. PHDC estimates may be higher in recent years and increasing over time due to greater ascertainment of patients using the public sector for HIV services. Overall, PHDC estimates appear to track Thembisa estimates closely in recent years, supporting the use of these routine individuated data for surveillance purposes in this setting. DHIS estimates include those currently on ART (excluding those interrupting ART) and trends may further represent changes in capturing of routine information on service-based registers or other contextual factors. While DHIS indicators are clearly defined, the exact question asked to ascertain current ART use at first antenatal visit may differ from clinician to clinician, resulting in variable patient interpretation and responses. Where data is not collected for research purposes it is difficult to determine how a question is asked. Data sources based on responses therefore may be uncertain, particularly with increasing ART coverage over time.

Population-wide datasets from the PHDC allow better discernment of current ART versus ever on ART and more detailed analysis such as district-level and age disaggregated temporal trends, which the antenatal survey is underpowered to generate, while the DHIS is limited to specific indicators. Age-group trends over time show an expected higher ART coverage prior to pregnancy in older age groups who have likely had more opportunities for HIV diagnosis and care than younger age groups (in whom HIV is likely to have been more recently acquired). District-level trends are in keeping with provincial trends, showing a high ART coverage during the antenatal period, with most districts above 90%. The lower and less precise coverage estimates in Central Karoo are reflective of the smaller population and lower HIV prevalence in this sparsely populated rural district. Other demographic characteristics associated with ART coverage could not be evaluated as these are not routinely captured in health information systems.

### Limitations

Routine data are subject to various caveats impacting both quality and completeness. Given the wide range of information systems used and the diverse staff members involved in data capturing, these data are subject to varying administrative and capturing errors. Completeness of data cannot be accurately quantified as demonstrated in the limited PHDC data available prior to 2015. Contextual factors at different facilities may further impact the quality of data - this includes changes in capturing approaches, staff training, staff turnover, and COVID-19 pandemic impacts. Ascertainment of both pregnancy and ART use is dependent on how electronic systems are used in different facilities. However, the PHDC approach to using multiple data sources to enumerate pregnancy and HIV ascertainment, may mitigate against gaps in single data sources such as DHIS. ART coverage ascertained from routine data is inferred through dispensing of ART from pharmacy. This does not necessarily translate to administration and adherence to ART.

This study highlights the challenges in comparing datasets with differing numerators and denominators, particularly with respect to treatment interruption. Routine data are limited to variables captured routinely. Since information specific to the first antenatal visit is not routinely captured in all public health facilities, it was not possible to distinguish between first and follow-up antenatal visits in the PHDC dataset. While first visit can be ascertained in DHIS, aggregated data are susceptible to double counting if the patient attends multiple antenatal clinics.

Detailed demographic and treatment characteristics are not captured, precluding a deeper understanding of epidemiological trends. DHIS consists of data elements for both national and provincial reporting purposes, some of which measure the same variable but are captured in different registers and by different staff members. As such, different data elements may be used for both numerator and denominator in order to calculate ART coverage. ART estimates may therefore vary depending on the data elements selected.

Since the national antenatal survey is underpowered for detailed estimates, routine data cannot be adequately validated at district, sub-district and age-group levels. Furthermore, the findings on self-reported ART use in surveys reflect findings in the last decade. We recommend repeating the validation to confirm if this concordance has persisted.

Lastly, it should be noted that while Thembisa estimates were used for comparison with antenatal survey and PHDC estimates, the Thembisa model is partially dependent on estimates from the antenatal survey for model calibration and assumptions on fertility are derived from the PHDC.

## Conclusion

Our study demonstrates the validity and utility of antenatal ART coverage using routine individuated data. While laboratory detection is the most accurate measure of ART use, this is neither affordable nor practical at a population level. Routine individuated data provide an efficient and less costly method of estimation compared to surveys and more accuracy and granularity than aggregate data. Trends in the Western Cape province are reassuring in that the large majority of HIV positive women are initiated on ART before or during pregnancy. This is supportive of the province soon meeting the third target of the Global Plan for elimination of mother-to-child transmission of 95% of HIV positive women initiating ART prior to or during pregnancy. Reasons for non-initiation during pregnancy require further investigation as this has serious implications for prevention of vertical transmission and child morbidity and mortality. Strengthening of routine information systems, including improved digitisation of point-of-care HIV testing and ascertainment of treatment interruption, in addition to being invaluable tools to directly support patient care, may allow more accurate epidemiological trend analysis for timely action and improved quality of HIV care.

## Data Availability

The PHDC and DHIS data underlying the results presented in the study are unconsented, de-identified routine service data housed by the Western Cape Department of Health. Release of these data to a public domain would violate the Data Access Policy of the Western Cape Department of Health. Data requests may be sent to the Western Cape Provincial Department of Health. The survey data are available from the National Department of Health, South Africa. Restrictions apply to the availability of these data, however these data may be requested from the National Department of Health, South Africa. Thembisa model data are available from https://www.thembisa.org/.

## Acknowledgements

The authors gratefully acknowledge the contributions of various collaborators, including Selamawit Woldesenbet, Emma Kalk, Florence Phelenyane, Adrian Puren, Kathryn Stinson, University of Cape Town Department of Pharmacology, the Western Cape Department of Health, the National Institute of Communicable Diseases, the South African National Department of Health and the Measurement and Surveillance of HIV Epidemics (MeSH) consortium.

## References

1. World Health Organization. Global guidelines on criteria and processes for validation: elimination of mother-to-child transmission of HIV and Syphilis monitoring. Int J Gynecol Obstet. 2014;143:155–6.

2. UNAIDS/World Health Organisation Working Group on Global HIV/AIDS and STI surveillance. Guidelines for assessing the utility of data from prevention of mother-to-child transmission (PMTCT) programmes for HIV sentinel surveillance among pregnant women. 2013; Available from: https://www.ncbi.nlm.nih.gov/books/NBK159001/pdf/Bookshelf_NBK159001.pdf (Accessed 30 January 2023)

3. Wessels J, Sherman G, Bamford L, Makua M, Ntloana M, Nuttall J, et al. The updated South African National Guideline for the Prevention of Mother to Child Transmission of Communicable Infections (2019). South Afr J HIV Med. 2020;21(1):1–8.

4. Mirkuzie AH, Sisay MM, Hinderaker SG, Moland KM, Mørkve O. Comparing HIV prevalence estimates from prevention of mother-to-child HIV transmission programme and the antenatal HIV surveillance in Addis Ababa. BMC Public Health. 2012;12(1).

5. Gouws E, Mishra V, Fowler TB. Comparison of adult HIV prevalence from national population-based surveys and antenatal clinic surveillance in countries with generalised epidemics: implications for calibrating surveillance data. Sex Transm Infect. 2008;84(Suppl I):i17–23.

6. Rehle T, Lazzari S, Dallabetta G, Asamoah-odei E, Rehle T. Second-generation HIV surveillance: better data for decision-making. Bull World Heal Organ. 2004;82:121–7.

7. Dee J, Calleja JMG, Marsh K, Zaidi I, Dee J. HIV Surveillance Among Pregnant Women Attending Antenatal Clinics : Evolution and Current Direction. JMIR Public Heal Surveill. 2017;3:1–8.

8. World Health Organization. Consolidated guidelines on HIV testing services [Internet]. 2015. Available from: https://www.who.int/hiv/pub/guidelines/hiv-testing-services/en/ (Accessed 30 January 2023)

9. Huerga H, Shiferie F, Grebe E, Giuliani R, Farhat J Ben, Van-Cutsem G, et al. A comparison of self-report and antiretroviral detection to inform estimates of antiretroviral therapy coverage, viral load suppression and HIV incidence in Kwazulu-Natal, South Africa. BMC Infect Dis. 2017;17(1):1–8.

10. Osler M, Hilderbrand K, Hennessey C, Arendse J, Goemaere E, Ford N, et al. A three-tier framework for monitoring antiretroviral therapy in high HIV burden settings. J Int AIDS Soc. 2014;17:18908.

11. Mofenson LM. Antiretroviral Therapy and Adverse Pregnancy Outcome: The Elephant in the Room? J Infect Dis [Internet]. 2015;jiv390. Available from: http://jid.oxfordjournals.org/lookup/doi/10.1093/infdis/jiv390

12. Jacob N. Western Cape Antenatal Survey Report 2014. Cape Town; 2014.

13. Woldesenbet S, Kufa T, Lombard C, Manda S, K A, M C, et al. The 2017 National Antenatal Sentinel HIV Survey, South Africa, National Department of Health. [Internet]. 2019. Available from: https://www.nicd.ac.za/wp-content/uploads/2019/07/Antenatal_survey-report_24July19.pdf (Accessed 30 January 2023)

14. Woldesenbet S, Lombard C, Manda S, Kufa T, Ayalew K, Cheyip M, et al. The 2019 National Antenatal Sentinel HIV Survey, South Africa [Internet]. 2021. Available from: https://www.nicd.ac.za/wp-content/uploads/2021/11/Antenatal-survey-2019-report_FINAL_27April21.pdf (Accessed 30 January 2023)

15. Johnson LF, Dorrington RE. Modelling the impact of HIV in South Africa’s provinces: 2023 update. Thembisa Version 4.6 [Internet]. 2023. Available from: https://www.thembisa.org/downloads (Accessed 30 July 2023)

16. Johnson LF, Mutemaringa T, Heekes A, Boulle A. Effect of HIV Infection and Antiretroviral Treatment on Pregnancy Rates in the Western Cape Province of South Africa. J Infect Dis. 2020;(July 2019):21–4.

17. Xia Y, Milwid RM, Godin A, Boily MC, Johnson LF, Marsh K, et al. Accuracy of self-reported HIV-testing history and awareness of HIV-positive status in four sub-Saharan African countries. Aids. 2021;35(3):503–10.

18. Mavhandu-Ramarumo LG, Tambe LAM, Matume ND, Katerere D, Bessong PO. Undisclosed exposure to antiretrovirals prior to treatment initiation: An exploratory analysis. South Afr J HIV Med. 2021;22(1):1–10.

19. Sithole N, Gunda R, Koole O, Krows M, Schaafsma T, Moshabela M, et al. Undisclosed Antiretroviral Therapy Use at Primary Health Care Clinics in Rural KwaZulu Natal South Africa: A DO-ART Trial Sub-study. AIDS Behav [Internet]. 2021;25(11):3695–703. Available from: 10.1007/s10461-021-03319-4

20. Kharsany ABM, Cawood C, Lewis L, Yende-Zuma N, Khanyile D, Puren A, et al. Trends in HIV Prevention, Treatment, and Incidence in a Hyperendemic Area of KwaZulu-Natal, South Africa. JAMA Netw Open. 2019;2(11):1–16.

21. Grobler A, Cawood C, Khanyile D, Puren A, Kharsany ABM. Progress of UNAIDS 90-90-90 targets in a district in KwaZulu-Natal, South Africa, with high HIV burden, in the HIPSS study: a household-based complex multilevel community survey. Lancet HIV. 2017;4(11):e505–13.

22. Jennifer TM-G, Julia R, Livia M, Mark S, Guy H, F.Xavier G-O, et al. ART denial: results of a home-based study to validate self-reported antiretroviral use in rural South Africa. AIDS Behav. 2019;23:2072–8.

23. Grabowski MK, Reynolds SJ, Kagaayi RH, Clarke W, Chang LC, Nakigozi G et. al. The validity of self-reported antiretroviral use in persons living with HIV: a population-based study. AIDS. 2018;32(3):363–9.

24. Kim A, Mukui I, Young P, Mirjahangir J, Mwanyumba S, Wamicwe J et. al. Undisclosed HIV infection and ART use in the Kenya AIDS Indicator Survey 2012: relevance to targets for HIV diagnosis and treatment in Kenya. AIDS. 2016;30(17):2685–95.

25. Jacob N, Rice B, Kalk E, Heekes A, Morgan J, Hargreaves J, et al. Utility of digitising point of care HIV test results to accurately measure, and improve performance towards, the UNAIDS 90-90-90 targets. PLoS One [Internet]. 2020;1–13. Available from: 10.1371/journal.pone.0235471

26. Astawesegn FH, Stulz V, Conroy E, Mannan H. Trends and effects of antiretroviral therapy coverage during pregnancy on mother - to - child transmission of HIV in Sub - Saharan Africa . Evidence from panel data analysis. BMC Infect Dis [Internet]. 2022;1–13. Available from: 10.1186/s12879-022-07119-6

